# Top-Down Genomic Surveillance Approach to Investigate the Genomic Epidemiology and Antibiotic Resistance Patterns of *Enterococcus faecium* Detected in Cancer Patients in Arkansas

**DOI:** 10.1101/2022.11.23.22282607

**Authors:** Zulema Udaondo, Kaleb Abram, Atul Kothari, Se-Ran Jun

## Abstract

Control of hospital-associated *Enterococcus faecium* infection is a strenuous task due to the difficulty of identifying transmission routes and the persistence of this nosocomial pathogen despite the implementation of infection control measures that have been successful with other important nosocomial pathogens. This study provides a comprehensive analysis of over one hundred *E. faecium* isolates collected from 66 cancer patients at the University of Arkansas for Medical Sciences (UAMS) between June, 2018 and May, 2019. In the top-down approach used in this study we employed, in addition to the 106 *E. faecium* UAMS isolates, a filtered set of 2167 *E. faecium* strains from the GenBank database to assess the current population structure of *E. faecium* species and, consequently, to identify the lineages associated with our clinical isolates. We then evaluated the antibiotic resistance and virulence profiles of hospital-associated strains from the species pool, focusing on antibiotics of last resort, in order to establish an updated classification of high-risk and multidrug-resistant nosocomial clones. Further investigation of the clinical isolates collected from UAMS patients using whole genome sequencing analytical methodologies (cgMLST, coreSNP and phylogenomics), with the addition of patient epidemiological data, revealed a polyclonal outbreak of three sequences types occurring simultaneously in different patient wards. The integration of genomic and epidemiological data collected from the patients increased our understanding of the relationships and transmission dynamics of the *E. faecium* isolates. Our study provides new insights into genomic surveillance of *E. faecium* to assist in monitoring and further limiting the spread of multidrug-resistant *E. faecium*.

## Introduction

Enterococci are common members of the gastrointestinal microbiota and other mucocutaneous membranes (1, 2). Although their virulence is low in healthy, immunocompetent individuals, enterococci have become the third leading cause of healthcare-associated infections in the United States (3). It has been reported that *Enterococcus* is one of the genera that most readily colonize the intestine after depletion of the indigenous intestinal microbiota, which occurs following therapeutic treatments, such as the use of antibiotics (4). More importantly, the development of *E. faecium* carriage in the gut has been highlighted as the most important risk factor for *E. faecium* bacteremia in immunocompromised patients (5, 6).

In the USA, recent reports indicate that 75% to 80% of the *E. faecium* infections reported in nosocomial settings are vancomycin-resistant *Enterococcus faecium* (VREfm) (7). VREfm strains that cause these infections in hospital settings are commonly resistant to all antienterococcal antibiotics that have been widely described (8, 9), making these infections often untreatable (10, 11). For that reason, VREfm is currently on the World Health Organization’s (WHO) list of high-priority pathogens for research and development of new antibiotics (12).

Daptomycin (DAP) is a cyclic lipopeptide antibiotic that has become the cornerstone in the thera-peutic treatment for severe infections caused by VREfm and methicillin-resistant *S. aureus* (MRSA) infections (13–15). As a last resort antibiotic, DAP is used in combination with other drugs or in sequential regimens after the failure of standard treatments (16) such as vancomycin (VAN) for infections caused by VREfm (17). Although DAP resistance in *E. faecium* is still rare, there has been an increase in recent years of treatment failures related to an emergence of DAP non-susceptible strains (18, 19). Major institutions and health care centers have reported that 20% to 30% of their VREfm isolates are DAP non-susceptible (DAP-NS) (20). Current knowledge suggests that the daptomycin non-susceptible (DAP-NS) phenotype is the result of a complex, multi-factorial mechanism which has its origin in specific mutations in the chromosome of the resistant strains (14). These mutations have been most consistently observed in two genes (*liaR* and *liaS)* that encode proteins related to phospholipid metabolism and cell envelope homeostasis (11, 21).

Whole-genome sequencing (WGS) methods have become crucial for outbreak and surveillance investigations in different settings due to their high resolution and backward compatibility with traditional methods such as multilocus sequence typing (MLST) (5, 22–25). These techniques have been successfully leveraged to investigate the transmission of *E. faecium* strains between livestock and humans (26), between multiple hospitals (27) and in hospital settings (19, 28). Thus, surveillance methods for infection prevention and control are rapidly moving toward integration of genomic and epidemiology data. The use of WGS-based surveillance methods of VREfm combined with epidemiological information is particularly relevant as there are a great number of epidemiologically unrelated cases that are caused by asymptomatic colonization, hampering the detection of smaller outbreaks (23) and acting as a reservoir for onward transmission (24).

In this study, we applied a top-down (from population genomics to molecular biology) genomic surveillance approach to investigate the genomic epidemiology and the antibiotic resistance patterns of nosocomial *E. faecium* strains, especially focusing on their VAN and DAP genotype. For this purpose, we employed all publicly available *E. faecium* genomes in GenBank database (at the time of the study) and a set of 106 *E. faecium* isolates from 66 cancer patients collected between June, 2018 and May, 2019 at the University of Arkansas for Medical Sciences (UAMS) (AR, USA). The goal of the present study is two-fold. The first aim is to leverage WGS and comparative genomics to expand the current knowledge of *E. faecium* distribution in its three clades (A1: hospital-associated, A2: environmental and animal-associated isolates, and human commensals that could cause occasional human infections, and B: community-associated) in order to analyze the antibiotic resistance patterns of hospital-associated *E. faecium* strains. The second aim is to describe the clinical and epidemiological traits of 106 clinical *E. faecium* strains isolated from UAMS cancer patients to better understand *E. faecium* epidemiology and transmission routes in the hospital setting.

## Results

### Genomic population structure of *Enterococcus faecium* species produces a seamless differentiation of three genetic lineages

The genome distances tree (Figure 1) of the entire set of genomes of *E. faecium* strains available to date (June, 2021) in the GenBank database (n=2167 *E. faecium* strains) and the set of UAMS isolates (n=106 *E. faecium* strains), constructed using Fast alignment-free computation of whole-genome Average Nucleotide Identity (FastANI, (29)), shows a clear division of the strains of this species into the two previously described groups, A and B, as well as a subsequent division of the A strains into clades A1 and A2 (Suppl. Table 1) (30, 31). Clade A1 (n=1384 total strains), formerly known as clonal complex 17 [CC17]), (32, 33), harbors strains obtained almost entirely from clinical and human sources (Figure 1, blue inner ring), with only one “avian” source sample, while clade A2 (n=695 strains) comprises *E. faecium* strains mainly from environmental and animal sources (Figure 1, orange inner ring). Clade B (n=194 strains) consists of strains that were isolated from environmental sources (mainly chickens and cattle). However, this clade also harbors a large number of strains classified as clinical strains from human samples (30) that are usually found in the gut microbiome of healthy individuals and rarely cause infections (34).

**Figure 1:**
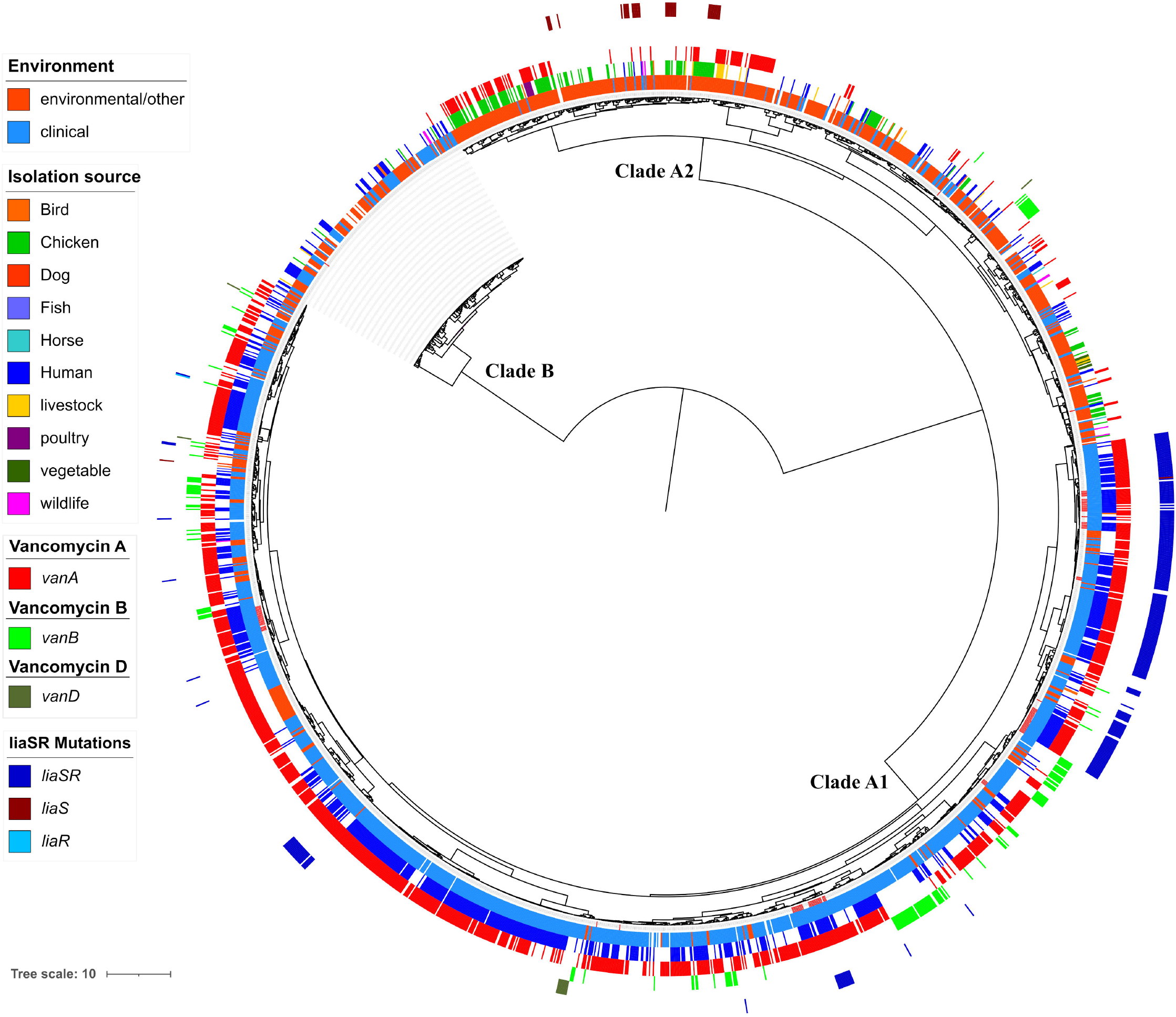
Pairwise genomic distance-based tree. Pairwise genomic distances were obtained using FastANI. The tree shows a clear division of the species into clades A1, A2 and B. The first color ring represents the environmental source (environment/clinical) from which the strain was isolated. The second color ring represents the isolation source. Presence of vancomycin resistance determinants, VanA, VanB and VanD are represented in rings 3 to 5. The outer ring represents the presence of mutations in *liaSR* genes, related to development of DAP non-susceptible phenotype.

According to the classification obtained in our analysis, the set 106 *E. faecium* strains isolated from UAMS patients have representatives in all three clades of *E. faecium* species with most of them (a total of 103 isolates) classified into clade A1. However, two of the isolates from two patients (UAMS_EF26 and UAMS_EF79) were classified into clade A2 and one as clade B (UAMS_EF24).

Isolate UAMS_EF26 was identified in the urine sample of a cancer patient at the emergency department and is genetically highly similar to *E. faecium* strain 306EA1 (ANI: 99.9003) that was isolated from the breast meat of a chicken processed in the state of Texas (USA) according to a study of enterococci associated with chicken meat from USA supermarkets (35). Strain 306EA1 was also classified as clade A2 in the study performed by Manson and colleagues (35). In the case of isolate UAMS_EF79, also obtained from the urine sample of a patient with cirrhosis in urinary analysis, this isolate was genetically closely related to *E. faecium* strain B3754 (ANI: 99.8334) that was isolated from the stool sample of a human neonate in the study developed by Liang and colleagues (36).

As previously stated, one isolate from the set of UAMS samples (UAMS_EF24), isolated from an outpatient in a routine urine analysis test, was identified within clade B. Isolate UAMS_EF24 is closely related to the strain *E. faecium* Com15 (ANI: 99.5602) which was obtained from the stool of a healthy individual (37) and classified as clade B in several studies (38, 39).

Our ANI-based lineage classification was further validated using the dataset provided by Lebreton *et al*., in 2013 (30) which identified the three *E. faecium* clades (A1, A2, and B) using a single nucleotide polymorphism (SNP)-based phylogenetic tree. We found only one discrepancy in the lineage classification between both datasets, that was the strain E. *faecium* E4452 which was allocated in clade A1 by the SNP-based lineage classification, but in clade A2 based on our ANI-based methodology. *E. faecium* strain E4452 is an ampicillin-resistant canine isolate assigned to ST266, a sequence type commonly found in dogs (40), thus fitting better with the characteristics of clade A2 (animal-associated).

Due to the growing number of VREfm infections in hospital settings that now represent up to 80% in some areas of the USA (7, 9), we studied the distribution of VAN resistance determinants in the context of the species, by mapping strains for which these determinants were found on the genome distances tree from Figure 1 (3rd - 5th rings from tree). This analysis unveiled that clade A1 harbored the highest number of VAN resistant strains with *vanA* (n=1053 clade A1 strains out of 1193 total strains with *vanA* operon identified in the species set), *vanB* (n=143 clade A1 strains out of 160 total strains with *vanB* operon identified in the species set) and *vanD* (n=8 clade A1 strains out of 10 total strains with *vanD* operon identified in the species set) genotypes (note that VanC-type resistance, is ordinarily intrinsic to species *Enterococcus gallinarum* and *Enterococcus casseliflavus*) *(41)*.

It has been reported that the use of the VAN analogues such as avoparcin, as a growth promoter on poultry, pig and cattle farms in Australia and the European Union (EU), promoted the proliferation of VAN resistance in farm settings (42–44). Our analysis shows that despite the ban of several antibiotics used as growth promoters, including avoparcin which has been banned since 1997 in the EU, over 22% of the strains from clade A2 still harbor antibiotic resistance determinants for VAN resistance genotypes *vanA* (n=139), *vanB* (n=17) or *vanD* (n=2) (note that the oldest strain used in our dataset was *released* in GenBank database in 2002 and the newest in 2021 (Suppl. Table 1).

As reported in several studies, LiaR (W73C) and LiaS (T120A) amino acid substitutions in proteins encoded by the LiaFSR (for lipid-II–interacting antibiotics) three-component cell envelope stress response system that regulates cell envelope integrity, are associated with the development of DAP non-susceptible phenotype in *E. faecium* (19, 21, 45). We queried our species set and identified 275 *E. faecium* strains (12% of the total species set) that harbored those specific co-mutations in *liaSR* genes (Figure 1, outer ring). These 275 strains were classified as clade A1 and a high proportion of them (n=243, 88%) also carried VAN resistance determinants. However, in the set of UAMS isolates, 63 out of 106 isolates (59%) had DAP non-susceptible mutations in *liaSR* genes, and 59 of them (94%) also carried the *vanA* gene cluster, thereby implying that the number of isolates with mutations related to the DAP non-susceptible phenotype is considerably overrepresented in the set of UAMS isolates.

### Resistome and multilocus sequence typing analysis of clade A1 strains indicate predominance of multidrug resistant clone ST80

The hospital-adapted lineage clade A1, which accounts for the vast majority of *E. faecium* invasive infections (5, 46), harbors the majority of VREfm strains including most of the UAMS isolates. To further characterize the resistome of clade A1 strains, we identified the antibiotic resistance determinants (ARDs) of the 1384 strains belonging to clade A1 using RGI tool against CARD database (47). A total of 48 ARDs (found in more than 1% of clade A1 strains) were annotated (Figure 2 and Suppl. Table 2). Moreover, using *in silico* MLST we classified a total of 1332 clade A1 strains into 74 sequence types (STs) that were interspersed throughout the tree shown in Figure 2A (inner ring).

**Figure 2:**
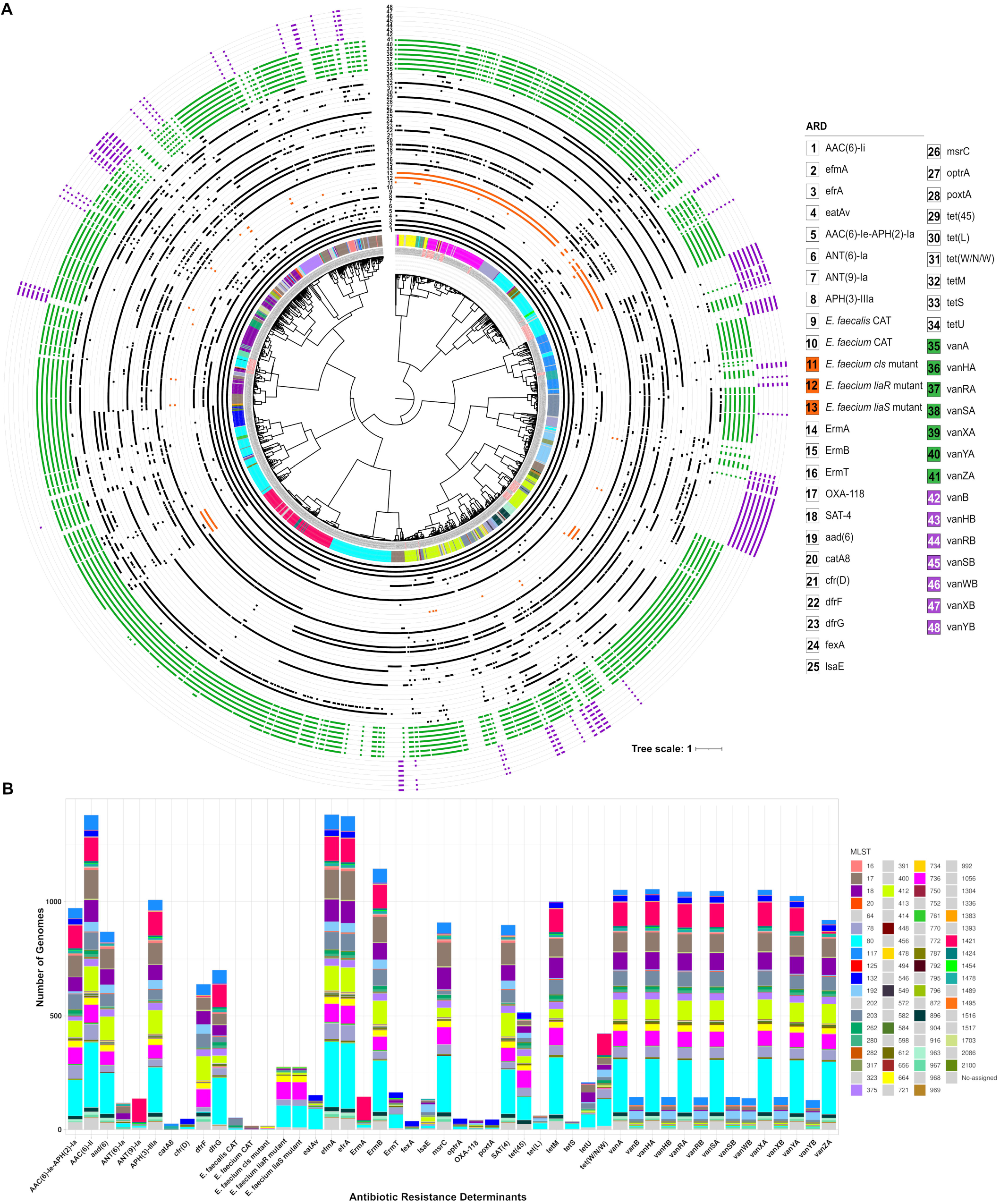
A. Pairwise genomic distance-based tree using ANI measurements of 1384 clade A1 *E. faecium* strains. The outer rings of the tree represent the resistome of each strain. The innermost ring represents sequence type affiliation. **B. Stacked barplot of ARDs identified in strains from clade A1 with MLST distribution**. A total of 48 non-redundant ARDs and 75 sequence types, were annotated for strains in clade A1.

The *in silico* MLST analysis revealed that the most predominant STs in clade A1 are ST80 (n=287 strains), followed by ST17, (n=126 strains), ST412 (n=107), ST1421 (n=104) and ST18 (n=97), which all together account for more than 54% of clade A1 strains. Four of these STs were also among the STs found with a higher level of multidrug resistance (in order of multidrug resistance level: ST17, ST80, ST18 and ST412). In the case of ST17, all 48 antibiotic resistance determinants were found in the resistome of strains belonging to this sequence type (Suppl. Table 2). However, two ARDs were not found among ST80 strains. Those two ARDs were chloramphenicol acetyltransferases (CAT): *catA8* and the *Enterococcus faecalis* CAT. In the case of ST18 strains, two ARDs, the rRNA-methylating enzyme codified by *cfr(D)* gene, that confers resistance to several classes of antimicrobial agents such as lincosamides, phenicols, and oxazolidinones, and the ATP-binding cassette protein that confers resistance to tetracyclines, phenicols, and oxazolidinones, *poxt(A)*, were not found in the resistome of the strains from this ST.

Notably, 12 out of 74 STs, do not harbor strains with *vanA* or *vanB* ARDs (these were, ST400, ST202, ST456, ST598, ST64, ST323, ST582, ST549, ST752, ST904, ST916, and ST413). Those 12 STs are among the least representative STs in clade A1 strains, with only one classified strain (two in case of ST549) and also being among the STs with the lowest number of annotated ARD. Dominant STs with the largest proportion of VREfm are ST1421 (99% of strains in this sequence type are VREfm), ST203 (85% of strains are VREfm), ST736 (99% of strains are VREfm) and ST80 (81% strains are VREfm).

Clade A1 strains are usually resistant to ampicillin and quinolone and have a higher rate of spontaneous mutation and recombination than clades A2 and B (33, 48). The stacked barplot in Figure 2B shows that the most abundant ARDs in clade A1 strains are the intrinsic *efmA* (present in 99.9% of clade A1 strains and confers resistance to macrolides and fluoroquinolones); and the chromosomalencoded *AAC(6’)-Ii* gene which was found in 99.8% strains and confers resistance to aminoglycosides. The *efrA* gene codifies for the EfrA efflux pump subunit and confers resistance to several antimicrobials such as rifamycin, fluoroquinolone and macrolides. It was reported for the first time in *E. faecium* species in 2013 (49), and was annotated in 99.4% clade A1 strains. The acquired ARD e*rmB*, which is induced by erythromycin and confers resistance to macrolides, lincosamides and streptogramins was annotated in 83% of clade A1 strains, followed by *vanA* ARD whose gene cluster was annotated in 76% of clade A1 strains.

The distribution of STs shown in the stacked barplot also indicates that there are some ARDs that can be found in virtually all STs (mainly those described in previous paragraph), but on the other hand, there are some ARDs that are preferably found in certain STs and not in others. For example, the chromosomally encoded dihydrofolate reductase, *dfrF* gene, that confers resistance to diaminopyrimidine antibiotic, was annotated in almost 50% of clade A1 strains, and is predominantly found in strains belonging to clones ST412 and ST736 (99% of strains belonging to these two STs harbor *dfrF* gene). By contrast, this ARD was found in only 1% of strains classified as ST80 and in 3% of ST736 strains. On the other hand, the dihydrofolate reductase *dfrG* gene, was annotated in 72% of ST80 strains and 95% of ST1421 strains but is only in a small percentage of ST412 and ST736 strains (29% and 2% of strains respectively) (Suppl. Table 2). In the same vein, strains with mutations in genes encoding the LiaFSR three-component regulatory cell envelope stress response system, associated with DAP non-susceptibility (21), were found in 100% of the strains from sequence types ST736, ST1454, and ST664. These co-mutations were also found in 67% of strains classified as ST280 and in 33% of strains classified as ST80 and ST734. Only a small proportion of clade A1 strains (1%), mainly classified as ST17, had mutations in the cardiolipin synthase (*cls*) gene which has also been associated with a predisposition to the non-susceptible phenotype to DAP (50).

Along with DAP, linezolid has become a last resort antibiotic for the treatment of VREfm infections since its introduction for clinical use in 2000 in the USA (51). In recent years, the increased use of linezolid has led to an exponential growth in the number of linezolid-resistant VREfm infections (52). Enterococcal linezolid resistance can be due to chromosomal mutations in the 23S rRNA gene and as well as by mutations in the genes encoding L3 and L4 ribosomal proteins (53). However, resistance can also be developed after the acquisition of the *optrA, poxtA* and *cfr-like* ARDs (53, 54). Genes *optrA, poxtA* and *cfrD* were annotated in a total number of 49, 45 and 48 clade A1 strains respectively. Analysis of the distribution of these three ARDs on the STs is very similar (Figure 2), and strains with these three ARDs primarily belonged to ST132. Thus, 92% of strains from ST132 harbor the 23S ribosomal RNA methyltransferase *cfrD* gene and more than 88% and 96% respectively, also harbor the *optrA* and the *poxtA* genes that codify for two ribosomal protection protein of the ABC-F protein families.

In silico virulence profiling of clade A1 strains using ABRicate v1.0.0 (Seemann T, *Abricate*, Github https://github.com/tseemann/abricate) and a virulence factor custom database (see methods section) unveiled a total of 4735 virulence factors in clade A1 strains codified by 12 genes (*ace, acm, ecbA, esp, gspG, hitB, hyl, ipxA, msbB, scm, sgrA* and *wecA*). The most predominant virulence factors found are the LPxTG-type surface exposed anchored proteins such as the collagenbinding microbial surface component Acm, which recognizes adhesive matrix molecules (MSCRAMMs), which was annotated in 91% of clade A1 strains; and the LPxTG surface adhesin, SgrA found in 86% of clade A1 strains. The SgrA virulence factor binds to fibrinogen and nidogen and is commonly implicated in biofilm formation. The sequence types with the highest number of virulence factor found were ST80 with a total of 916 annotated virulence factors (Suppl. Table 3), ST1421 with a total of 439 virulence factors and ST17 and ST18, with 427 and 320 annotated virulence factors respectively. Nevertheless, the profile of virulence factor abundance varies among the four STs (Suppl. Figure 1). For example, although members of ST1421 were not among the most highly multidrug resistant strains of the analysis, this sequence type has the highest percentage of strains harboring most of the virulence factors annotated in clade A1 strains. Thus, 100% of the ST1421 clade A1 strains harbored the virulence factors Acm (non-fimbrial adhesin) and EcbA (non-fimbrial adhesin) in their genome; 99% of ST1421 strains had the virulence factor SgrA (non-fimbrial adhesin) and 98% harbored Hyl (hyalurodinase) virulence factor (Suppl. Table 3).

### Genomic epidemiology of 106 *E. faecium* isolates from cancer patients in Arkansas reveals persistence of clones in urine samples

In this study, we retrospectively sequenced the whole genome of 106 *E. faecium* isolates which belonged to a total of 66 cancer patients ranging in age from 23 to 87 years. All patients presented underlying medical conditions, and 17 of them had more than one sample positive for *E. faecium*, with a range of 2 to 12 isolates per patient (Suppl. Table 4).

Single nucleotide polymorphism (SNP)-based methods are widely used for hospital outbreak investigation as they retain higher resolution for tracking transmissions between patients (55). Thus, we constructed a pairwise coreSNP distance matrix with the set of UAMS isolates. The coreSNP distance matrix was plotted as a clustered heatmap that shows several clusters of highly genetically related isolates differing by less than 50 SNPs (Figure 3). The isolates within these clusters belong to the same sequence type and are, in almost all cases, independent of isolation source (blood or urine). The clusters with less than 50 SNPs were also confirmed by core genome MLST (cgMLST) analysis using the official *E. faecium* scheme with 1423 core genes (56). The number of cgMLST allelic differences between all pairs of isolates was calculated and compared with the coreSNP distance matrix. Isolates with at least 5 allelic differences were classified into different complex types (CTs) yielding a total of 89 CTs for all 106 isolates (Figure 4).

**Figure 3:**
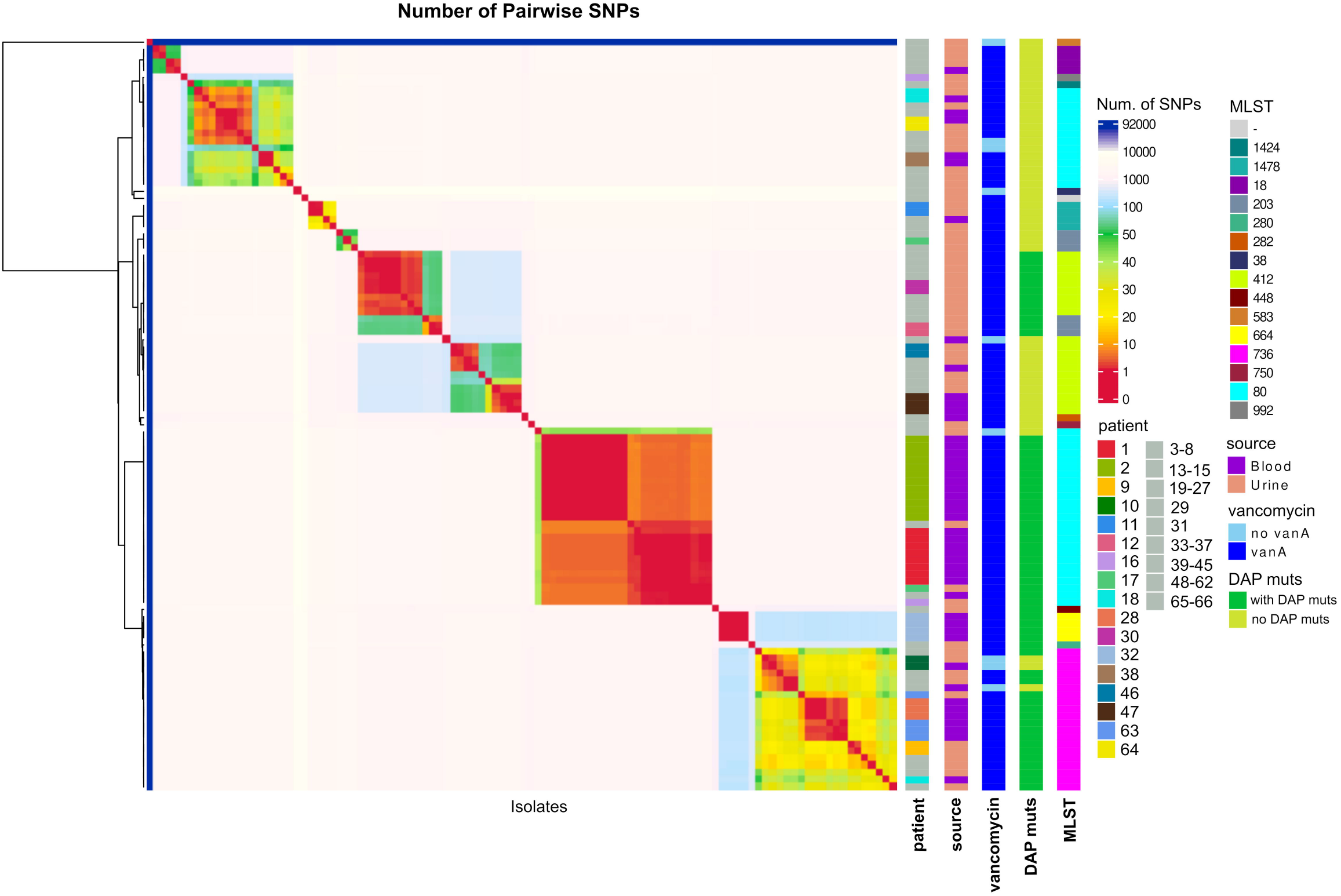
Pairwise coreSNP heatmap of 106 *E. faecium* isolates from 66 cancer patients. The heatmap was colored using pairwise coreSNP distance. Warmer colors in the heatmap represent clusters of *E. faecium* clones. Metadata including patient number, sequence type, isolation source, presence of *vanA* cluster and presence of daptomycin non-susceptibility related mutations, was annotated. Patients with only one isolate are shown in grey color.

**Figure 4:**
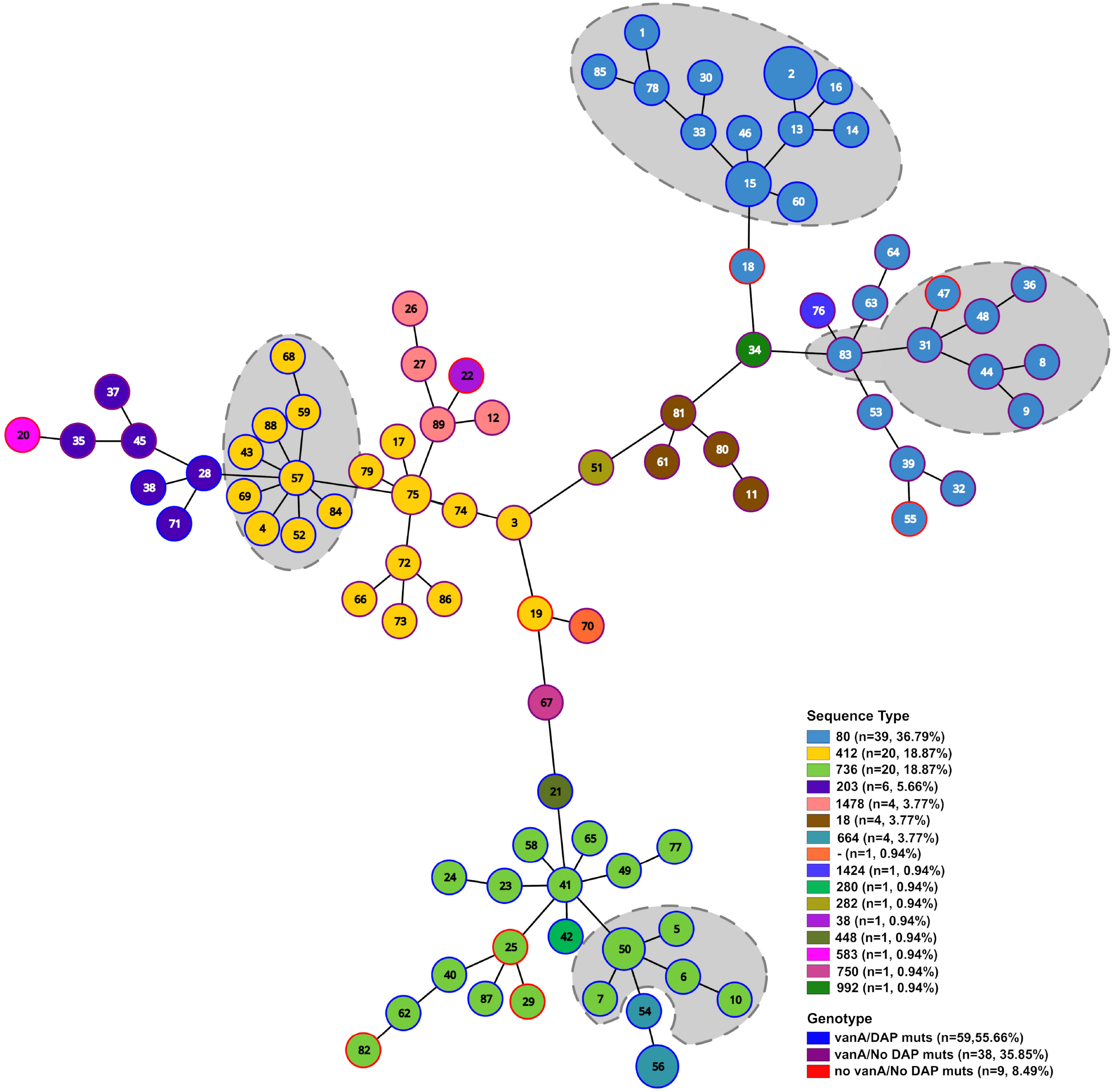
Minimum spanning tree of 106 *E. faecium* isolates from cancer patients. Numbers in node represent the cgMLST classification. Node size represents number of strain for each cgMLST. Node color represents sequence type. Different combinations of presence/absence of vanA cluster and DAP non-susceptibility related mutations are shown in the outer ring of each node in blue, purple and red color. Percentage in parentheses represents the relative abundance of strains in that category. Grey background delineates isolates that were detected as part of clonal cluster with a coreSNP threshold of 15.

Based on previous VREfm surveillance studies, patients with isolates differing by less than 15 SNPs are considered to be closely related genetically and within the limits of putative transmission (23, 57). Using the threshold of 15 pairwise coreSNPs, we identified four major *E. faecium* clonal clusters prevalent among UAMS samples that were collected at different time points throughout one year (Suppl. Table 4 and Suppl. Table 5). Isolates belonging to these four clonal clusters had 14 or less cgMLST allelic differences. Manual inspection of the coreSNPs found between strains belonging to the four clonal clusters, showed a limited number of recombination events (0 to 2 in the set of strains) that were not taken into account in our threshold.

The four major clonal clusters were further investigated using a maximum likelihood tree of 103 clade A1 isolates which was rooted with the two UAMS isolates (UAMSEF_26 and UAMSEF_79) placed in clade A2 (Figure 5, Suppl. Figure 2 to 6). The main epidemiological characteristics of the four main clonal clusters of this polyclonal outbreak are described below.

**Figure 5:**
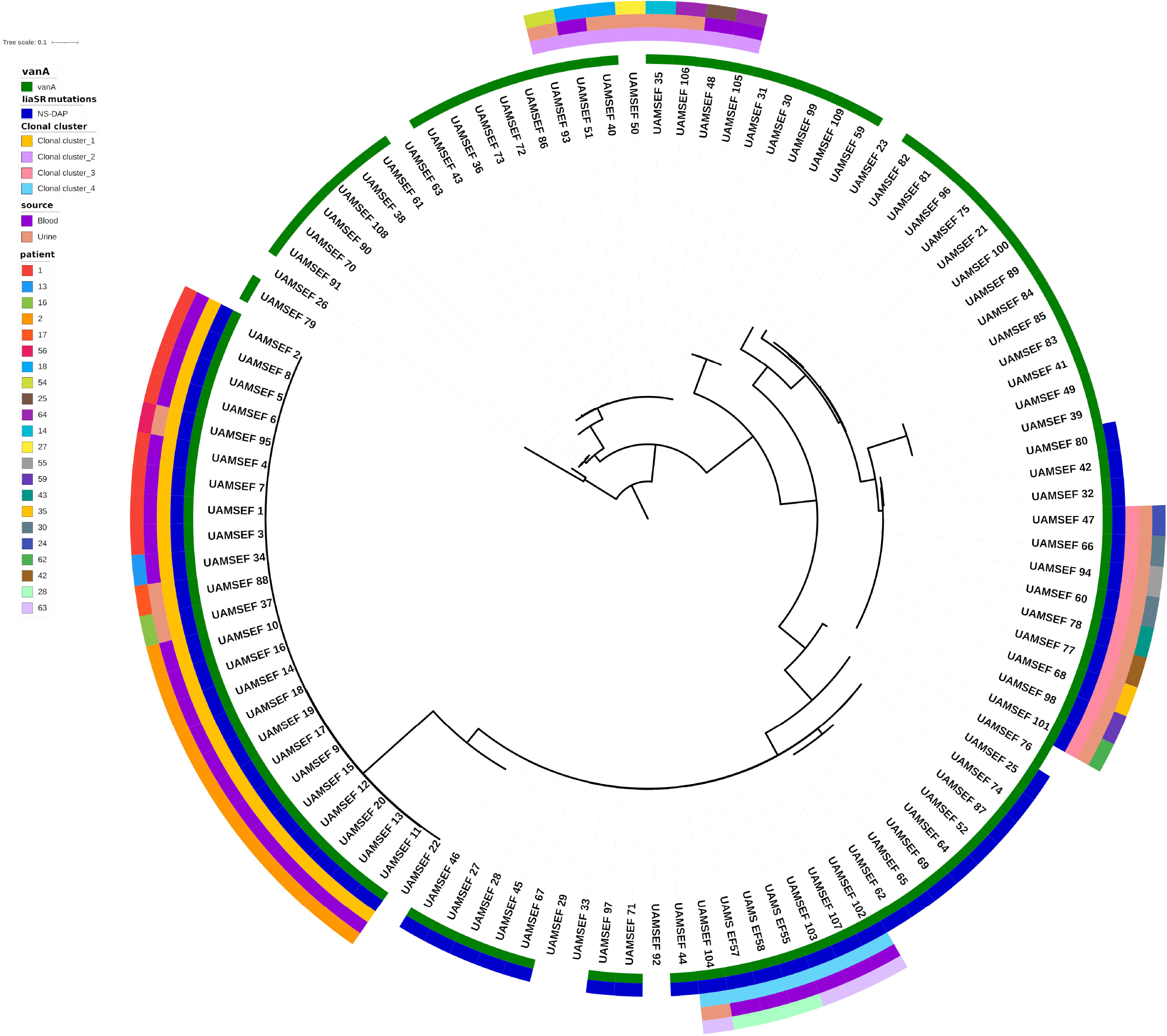
Maximum likelihood phylogenetic tree of 105 clade A1 and A2 *E. faecium* isolates. Isolates belonging to each of the four clonal clusters are marked in the inner ring of the tree. Presence of *vanA* cluster of genes conferring resistance to vancomycin and mutations in *liaSR* genes related to daptomycin non-susceptible phenotype are represented in rings 2 and 3 respectively.

#### Clonal cluster_1 of 24 VREfm ST80 isolates and with DAP non-susceptible mutations

This cluster involved isolates that were obtained from six different patients with different underlying hematologic malignancies (Suppl. Table 5). Four of these inpatients shared the same bed unit at different time points. Patient 17 was at the time of isolate collection, an outpatient of the UAMS cancer infusion center with a history of prior admissions to the hospital with overleaping dates with other patients at the time of higher prevalence of this clone at the hospital in which the same clone was found in at least two other hospitalized patients concurrently (Suppl. Table 3 and 5). Most of the isolates from this cluster belong to two patients (isolates UAMS_EF1 to 8 from patient 1, and UAMS_EF9 to 20 from patient 2) whose specific cases were previously described by us (19) due to the observed development of the DAP non-susceptible phenotype during treatment with DAP of these two patients. According to the phylogenetic analysis (Figure 5 and Suppl. Figure 2 and 3), isolates UAMSEF_34, UAMSEF_37, and UAMSEF_88 originated from patient’s 1 isolate lineage. Remarkably, isolates UAMSEF_88 and UAMSEF_95, both isolated from urine samples of two different patients, and collected three and four months after the discharge of the last patient registered in the hospital and from whom strains belonging to this cluster were isolated. The integration of genomic and epidemiologic data, indicates an extended prevalence of this clone in the gastrointestinal or urinary tract of the patients several months after the time of infection.

#### Clonal cluster_2 of eight ST80 isolates with diverse vancomycin genotypes

This cluster comprises isolates found in six patients that had underlying conditions of differing nature, including carcinoma, cholangiocarcinoma, and acute lymphoblastic leukemia. Most of the patients from this cluster had overlapping hospital stays (Suppl. Table 5). However, all patients were hosted in different bed units, indicating that *E. faecium* clones may have spread throughout different wards. None of the isolates from this cluster harbored DAP non-susceptible related mutations in *liaSR* genes and one isolate (UAMSEF_50) even showed a vancomycin-susceptible phenotype (*vanA* cluster was also not found in this isolate). According to cgMLST, coreSNP, ANI and phylogenetic analyses, isolate UAMSEF_50 is most closely related to UAMSEF_35. As in the previous clonal cluster_1, we observed the presence of isolates from urine samples which harbored very high genetic similarity to isolates collected from patients who had been discharged several months earlier (UAMSEF_93, UAMSEF_106) indicating a possible long-term prevalence of this clone in outpatients.

#### Clonal cluster_3 of nine ST412 VREfm and with DAP non-susceptible mutations

This outbreak involved isolates obtained from urine samples from eight different patients that had underlying conditions of differing nature, but mainly related to hematological malignancies. Patients remained in different bed units in most cases. Interestingly, although all isolates in this outbreak have the *vanA* gene cluster responsible for vancomycin resistant phenotype, one of the isolates (UAMS_EF47 from patient 24) had a vancomycin-susceptible phenotype. This isolate was collected at the emergency department during the first day of admission of patient 24. Since this patient was not hospitalized at the time of sample collection, the clone identified in the urine sample was marked as *community acquired*. However, examination of the epidemiological data revealed that patient 24 had been previously admitted to the hospital due to pancreas and kidney transplantation and was on prophylactic antibiotic treatment, indicating a possible long-term prevalence of this clone in this patient.

#### Clonal cluster_4 of six ST736 VREfm isolates with DAP non-susceptible mutations

Two patients diagnosed with acute myeloid leukemia and end-stage renal disease comprised this cluster. Both patients remained in different bed units, and their discharge and admission occurred on consecutive days.

## Discussion

*E. faecium* causes infections mainly in immunocompromised patients posing a serious threat to patients in intensive care units, burn patients, oncology treatment, and organ transplantation (58) which makes surveillance of this microorganisms in hospital settings extremely important.

In order to understand the distribution of a large collection of *E. faecium* clinical isolates collected from cancer patients at UAMS, we first leveraged all the genomes available in GenBank database (as of June 2021) and constructed the genomic population structure of *E. faecium* species using pairwise ANI distances. The subdivision of *E. faecium* strains into clades A1 and A2 has been argued in previous studies (46). However, the use of a larger data set with strains from broader geographical and ecological origins in combination with WGS-based methodologies, illustrated a strikingly clear distribution of the three clades of *E. faecium* species (Figure 1). Clade A1 strains dominate the hospital population of disease-causing *E. faecium* (59). Thus, most of the isolates obtained in our surveillance study on cancer patients at high risk were classified as members of *E. faecium* clade A1 (59).

Although clade A2 strains were mainly isolated from animal sources, we identified 54 strains within this clade A2 obtained from human samples. Most A2 strains from humans, as in the case of strain UAMS_EF26 from the set of UAMS samples, have a high sequence similarity to strains obtained from animal sources. Evidence of transient intestinal colonization by *E. faecium* strains of animal origin, capable of transferring mobile elements with resistance genes to the host microbiota, has been provided in other studies (60–62), imposing a potential threat to patients with weakened immunity and whose gut microbiome is under selective pressure from antibiotics.

We then focused our analysis on the clade A1 strains. The *in silico* MLST classification of clade A1 strains resulted in 74 STs. The ability of MLST analysis to discern between *E. faecium* strains has been discussed in multiple studies reaching different conclusions due to the high frequency of recombination events in this species that can affect several of the genes employed in MLST analysis (46). However, due to the rapid and unambiguous procedure, MLST is still widely used for global and long-term epidemiology of *E. faecium* and other bacterial species (63, 64). In line with previous studies, the current *E. faecium* MLST scheme does not accurately reflect the population structure of the clade A1 *E. faeciu*m strains (Figure 2A) (65). However, it seems to maintain a certain degree of correspondence with several monophyletic groups observed in the genomic distance tree of clade A1 strains (Figure 2A). Therefore, for genomic surveillance purposes, early identification of *E. faecium* isolates by their STs may still be useful as an initial preventive infection control measure, especially if there is prior knowledge of the resulting sequence type as belonging to a multidrug-resistant and pathogenic lineage (58).

Due to the above, we further investigated the possible relationships between the antibiotic resistance determinants, virulence factors, and the sequence types of clade A1 strains. Our results showed that currently, the most predominant sequence type in clade A1 strains is ST80 (n=287 strains), followed by ST17 (n=126 strains), ST412 (n=107), ST1421 (n=104) and ST18 (n=97) which all together account for more than 54 % of clade A1 strains. Among these sequence types were the STs found with the highest level of multidrug resistance (ST17, ST80, ST18 and ST412 ordered by multidrug resistance level). The sequence type ST1421 had the highest percentage of strains harboring most of the different classes of virulence factors annotated in clade A1 strains. This information follows the trend already observed in this species of emergence of highly hospital-adapted *E. faecium* clones with increased development of multidrug resistance which is related to the broad ability of *E. faecium* to survive in hostile antimicrobial-rich environments (66, 67). The process by which multidrug resistant clones develop is known as “genetic capitalism” (68) which states that an increase in fitness through the accumulation of adaptive elements, such as ARDs, by specific clones in selective environments, further increases the probability of acquiring more adaptive elements, leading to the emergence of high-risk multidrug-resistant clones as in the case of clade A strains (53).

Despite that vancomycin resistance determinants are harbored by the majority of strains and sequence types of clade A1 strains (Figure 2), ARDs linked to two important last-line antibiotics (such as linezolid and daptomycin) against VREfm infections, still seems to be limited to a small percentage of clade A1 strains that belong to different sequence types. Most strains with ARDs for linezolid (*optrA, poxtA* and *cfrD* genes) belonged to ST132 and, to a lesser extent, ST612 (Suppl. Table 2). On the other hand, most strains with mutations in the *liaFSR* operon associated with the development of non-susceptible phenotype to DAP belong to ST736 and ST664 (Suppl. Table 2).

Previous genomic studies of clade A1 *E. faecium* strains have revealed substantial levels of genome plasticity (67) and have provided evidence of polyclonal outbreaks in individual hospitals (59). The use of WGS-based approaches such as coreSNP (Figure 3), cgMLST (Figure 4), and phylogenetic analysis (Figure 5, Suppl. Figures 2-6) in combination with epidemiological data from patients, allowed us to identify a total of four *E. faecium* clonal clusters that simultaneously occurred at UAMS during the one-year sample collection period. Isolates from these four clusters belonged to three different sequence types, which are among the previously identified multidrug resistant clade A1 sequence types, ST80, ST412 and ST736. Most of the findings described in this study strongly suggest nosocomial transmission of the *E. faecium* clones within and between hospital wards, however we also found highly genetically related clones that persisted over time and were isolated from urine samples of immunocompromised patients that came to the hospital for routinary visits (Suppl. Table 4, Suppl. Table 5). Long-term persistence of multidrug-resistant enterococci clones in the gut of patients affected by systemic antimicrobial treatments has been documented (69, 70), as well as the existence of symptomatic and asymptomatic recurrent urinary tract infections caused by other species such as *Escherichia coli* (71, 72). Thus, the antimicrobial therapy which patients with chronic and hemato-oncological diseases are subjected to, as is the case of the patients in this study, may act as a selective and driving factor that promotes persistent enterococcal colonization through the inhibition of the autochthonous flora (70), and also through the induction of the expression of factors, other than ARDs, that could promote adherence to the intestinal epithelial lining and biofilm formation (69, 73). In the case of the UAMS dataset, several isolates from urine samples belonging to clusters of the outbreak described in our analysis were flagged as “community-acquired” because the isolate was identified in a routine analysis of a non-hospitalized patient, which might prevent early infection control management in these cases.

In conclusion, in this study we have performed a retrospective investigation of *E. faecium* isolates detected in blood and urine samples from high-risk patients who test positive for *E. faecium* using WGS analytical techniques in combination with clinical and epidemiological data in order to assess the genomic diversity of *E. faecium* isolates in the context of the species. Our findings demonstrate that the top-down genomic surveillance approach utilized in this study could accurately resolve a polyclonal outbreak of VREfm at UAMS. The addition of epidemiological data on high-risk patients who test positive for *E. faecium* along with ongoing genomic surveillance may be key to predict the carriage of persistent *E. faecium* clones, allowing early activation of infection control measures such as screening and contact precautions among such patients.

## Methods

### Study Design

The present study involved adult patients, men and/or women, ages ranging from 18 to 95 years old inclusive, undergoing treatment at the University of Arkansas for Medical Sciences (UAMS) (AR, USA), who develop a clinical infection (bloodstream or urinary tract infection as determined by blood or urine culture) with *Enterococcus faecium*. Patients who had blood or urine cultures positive for *E. faecium* (by standard clinical lab testing) were included in the study. Deidentified clinical and epidemiologic data was collected for the study participants. Clinical data includes outcome data related to hospital length of stay, length of ICU stay, hospital unit, duration of bacteremia, antimicrobial administered, 30 and 90 day mortality and development of Graft versus host disease. The study was approved by the Institutional Review Board of UAMS (IRB No. 228137).

### Bacterial isolates

A total of 106 *E. faecium* isolates were identified from positive blood and urine cultures from 66 patients. Samples collected were grown on blood cultures and processed on the BacT/ALERT 3D (bioMérieux) system. Positive blood cultures were then subcultured on blood agar plates. Isolated colonies were used for identification and susceptibility testing using the Vitek MS and Vitek 2 systems.

### DNA extraction and quantification

Microbial DNA was extracted from pure growth of VREfm. Isolated colonies on the blood agar plates were picked and re-suspended into a DNA/RNA Shield Collection and Lysis Tube (Zymo Research, Irvine, CA). Genomic DNA was extracted from the tube using Quick-DNA Fungal/Bacterial kit (Zymo Research, Irvine, CA). The purity of extracted DNA was determined using a NanoDrop Spectrophotometer by measuring the A260/280 and A260/230 ratios. DNA integrity and quantity were determined using an Agilent 2200 TapeStation and Qubit® 3.0 Assay, respectively.

### Vancomycin and daptomycin susceptibility testing

Vancomycin resistance was confirmed using E-tests (bioMérieux). Urine cultures were processed similarly using blood agar plates for isolation of colonies. Antimicrobial susceptibility test results were interpreted using the M100 CLSI standards (74).

### Illumina sequencing and Genome Assembly

Paired-end 150bp libraries were constructed using the KAPA HyperPlus kit (Roche) with enzymatic fragmentation for 10 min. The resulting genomic libraries of the *E. faecium* isolates were sequenced using the Illumina NextSeq 550 platform at the UAMS Myeloma Center. Adapters were trimmed using fastp v0.19.5 (75) with default settings. Trimmomatic v0.38 (76) was used to remove poor-quality reads, with the following parameters: HEADCROP:15 LEADING:20 TRAILING:20 SLIDINGWINDOW:5:20 MINLEN:50. Quality of pre- and post-processed reads was assessed with FastQC tool v0.11.8 (77). The resulting high-quality reads were assembled *de novo* using SPAdes v3.13.0 (78) with settings, “error-correction” and “careful”, k-mer sizes of 21, 33, 55 and 77 and minimum contig size of 500 bp. Draft genomes were quality-checked using default settings in QUAST v5.0.2 (79). Genome sequences were submitted for annotation to the NCBI Prokaryotic Genome Annotation Pipeline (PGAP) (80) using the default parameters.

### MLST and cgMLST scheme creation

MLST classification of the 1384 clade A1 strains was done using “mlst” software from T. Seemann (https://github.com/tseemann/mlst) which incorporates components of the PubMLST database (https://pubmlst.org/) developed by Keith Jolley (81) and sited at the University of Oxford. cgMLST classification of the set of 106 UAMS *E. faecium* isolates was obtained using chewBBACA suite v2.0.16 (82). The minimum spanning tree was plotted using PHYLOVIZ v2.0 applying goeBURST clustering algorithm (83).

### coreSNP analysis

Mapping of SNP calling analyses were done using Snippy v4.6 https://github.com/tseemann/snippy. Pairwise SNPs were calculated in R using harrietr (v0.2.3, https://github.com/andersgs/harrietr) and coreSNP alignments from Snippy without masking prophage or recombination regions as considered best for accurate and consistent MDR organism transmission inference, when using core genome alignments and SNP thresholds (57, 65). The data from the coreSNP analysis was represented as a clustered heatmap using R and ComplexHeatmap v2.12.1 library (84).

### FastANI tree

ANI measurements were obtained using FastANI v1.33 software (29) with a k-mer length of 21 and were used to construct a distance matrix (100 - ANI similarity measure). Distance-based trees were built using ape R package (85) and ward.D2 clustering algorithm. Trees were visualized using the Interactive Tree of Life, iTol v6.2 (86).

### Antibiotic resistance determinants and virulence factors annotation

Antibiotic resistance determinants were predicted using the Resistance Gene Identifier (RGI) v5.2.0 against the Comprehensive Antibiotic Resistance Database (CARD) database v3.1.1 (47), and virulence factors were identified using the ABRicate v1.0.0 screening tool using a custom database of virulence factors downloaded from NCBI nucleotide database and the virulence factor database (VFDB) accessed on August 26, 2022 (87, 88). Virulence factors with greater than 80% sequence coverage and sequence identity were annotated in the assemblies of clade A1 strains.

### Phylogenetic tree

Alleles obtained from the cgMLST analysis and present in all strains (n=1266 alleles) were extracted and individually aligned using MAFF v7.475 (89) with options --maxiterate 1000 --globalpair; using the algorithm G-INS-i for alignment strategy which assumes that entire regions can be aligned and tries to align them globally using the Needleman-Wunsch algorithm. Alignments were trimmed using ClipKIT v1.30 (90) and concatenated to be used as input for IQ-TREE v2.2.0.3 to construct a phylogenetic tree using a maximum likelihood approach (91). ModelFinder (92) was used to find the best model for each partition in IQ-TREE (-m MFP+MERGE). For the tree reconstruction, 1000 ultrafast bootstraps (-B 1000) (93) were used to evaluate the nodal support. The tree was rooted using clade A2 isolates.

## Data Availability

Raw sequencing data, assembly and functional annotations for the isolates used in this study are available under the BioProject accession numbers, PRJNA518133, PRJNA735268 and PRJNA520878, of NCBI database.

## Acknowledgments

Data processing for this work was performed in part using the High-Performance Computing Center which is funded through multiple National Science Foundation grants and the Arkansas Economic Development Commission.

## Ethics approval and consent to participate

This study was approved by the Institutional Review Board of University of Arkansas for Medical Sciences (IRB No. 228137).

## Author contributions

Conceptualization: S.J. Resources: A.K. Funding acquisition: S.J. Supervision: S.J. Methodology: Z.U. Formal analysis: Z.U. and K.Z.A. Investigation: Z.U. Visualization and writing – original draft: Z.U. Writing – review and editing and validation: Z.U., A.K., K.Z.A. and S.J.

## Supplementary Information

**Supp. Table 1**. List of all 2273 strains used the study with relevant associated metadata.

**Suppl. Table 2**. Number (sheet 1) and frequencies (sheet 2) of antibiotic resistance determinants identified in 1384 clade A1 *E. faecium* strains by MLST. The symbol “-” is used for strains without sequence type.

**Suppl. Table 3**. List of virulence factors identified in 1384 clade A1 *E. faecium* strains by MLST.

**Suppl. Table 4:** Outbreak information. Characteristics for each of the clonal clusters identified in the polyclonal outbreak is highlighted in this table.

**Suppl. Table 5**. Gantt chart of the polyclonal outbreak. The hospitalization date and the ward location in the hospital at the time of isolate collection are represented.

**Suppl. Figure 1:** MLST and virulence factor intersection plot. Numbers above the bars represent the number of sequence types for each set of virulence factors (indicated by black circles in the intersection plot).

**Suppl. Figure 2:** Maximum likelihood phylogenetic tree of 105 clade A1 and A2 *E. faecium* isolates. Isolates belonging to each of the four detected clonal clusters are marked in the inner ring of the tree. Presence of *vanA* cluster of genes conferring resistance to vancomycin and mutations in *liaSR* genes related to daptomycin non-susceptible phenotype are represented in the second and third bar respectively. Bootstrap values are represented in the tree branches.

**Suppl. Figure 3:** Pruned maximum likelihood phylogenetic tree of clade A1 and A2 *E. faecium* isolates which contains strains from clonal cluster 1. Presence of *vanA* cluster of genes conferring resistance to vancomycin and mutations in *liaSR* genes related to daptomycin non-susceptible phenotype are represented in the second and third bar respectively. Bootstrap values are represented in the tree branches.

**Suppl. Figure 4:** Pruned maximum likelihood phylogenetic tree of clade A1 and A2 *E. faecium* isolates which contains isolates from clonal cluster 2. Presence of *vanA* cluster of genes conferring resistance to vancomycin and mutations in *liaSR* genes related to daptomycin non-susceptible phenotype are represented in the second and third bar respectively. Bootstrap values are represented in the tree branches.

**Suppl. Figure 5:** Pruned maximum likelihood phylogenetic tree of clade A1 and A2 *E. faecium* isolates which contains isolates from clonal cluster 3. Presence of *vanA* cluster of genes conferring resistance to vancomycin and mutations in *liaSR* genes related to daptomycin non-susceptible phenotype are represented in the second and third bar respectively. Bootstrap values are represented in the tree branches.

**Suppl. Figure 6:** Pruned maximum likelihood phylogenetic tree of clade A1 and A2 *E. faecium* isolates which contains isolates from clonal cluster 4. Presence of *vanA* cluster of genes conferring resistance to vancomycin and mutations in *liaSR* genes related to daptomycin non-susceptible phenotype are represented in the second and third bar respectively. Bootstrap values are represented in the tree branches.

