## Supplementary figures and images for "Top-Down Genomic Surveillance Approach to Investigate the Genomic Epidemiology and Antibiotic Resistance Patterns of *Enterococcus faecium* Detected in Cancer Patients in Arkansas"

### Suppl. Fig2

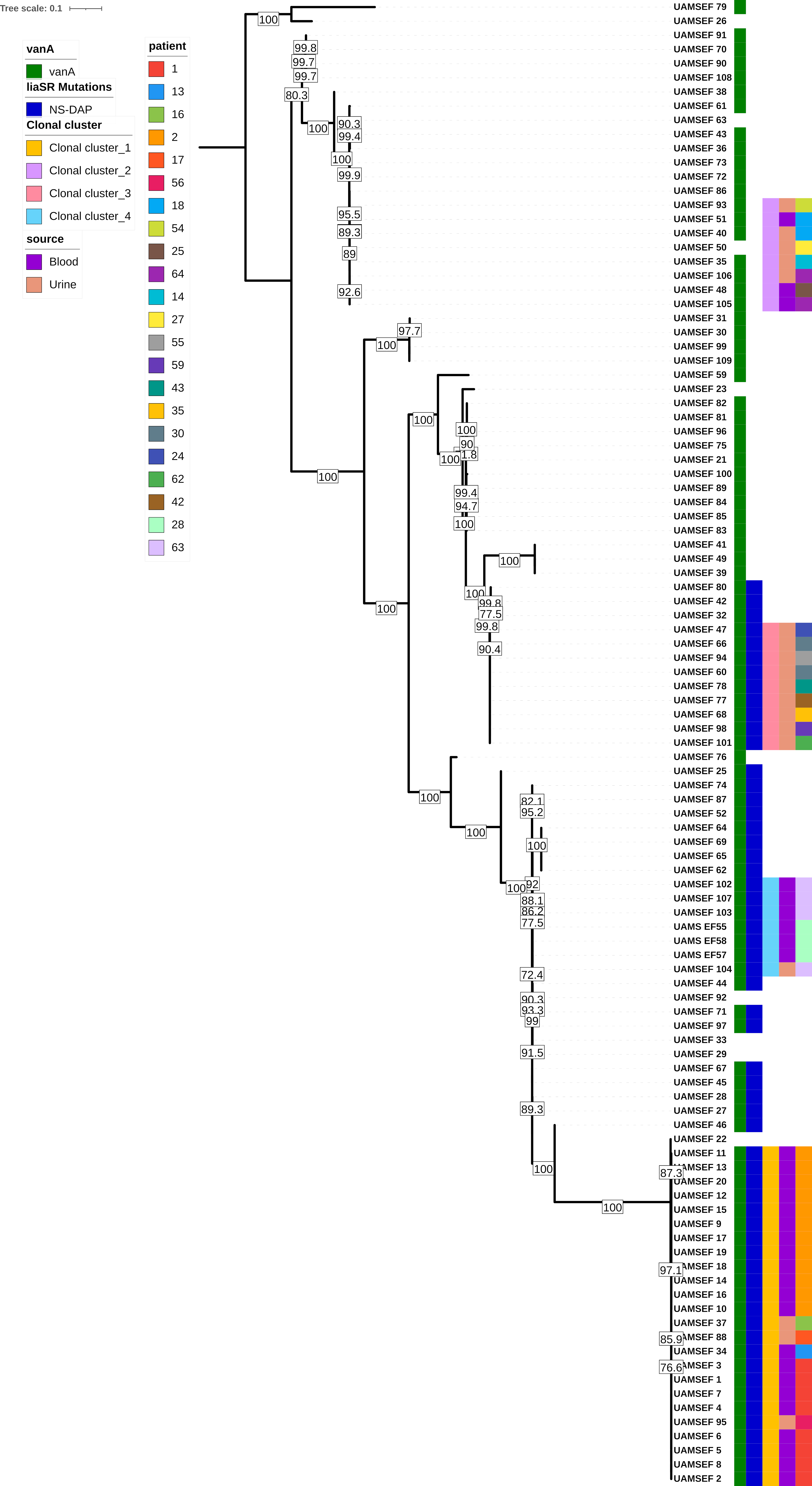

### Suppl. Fig3

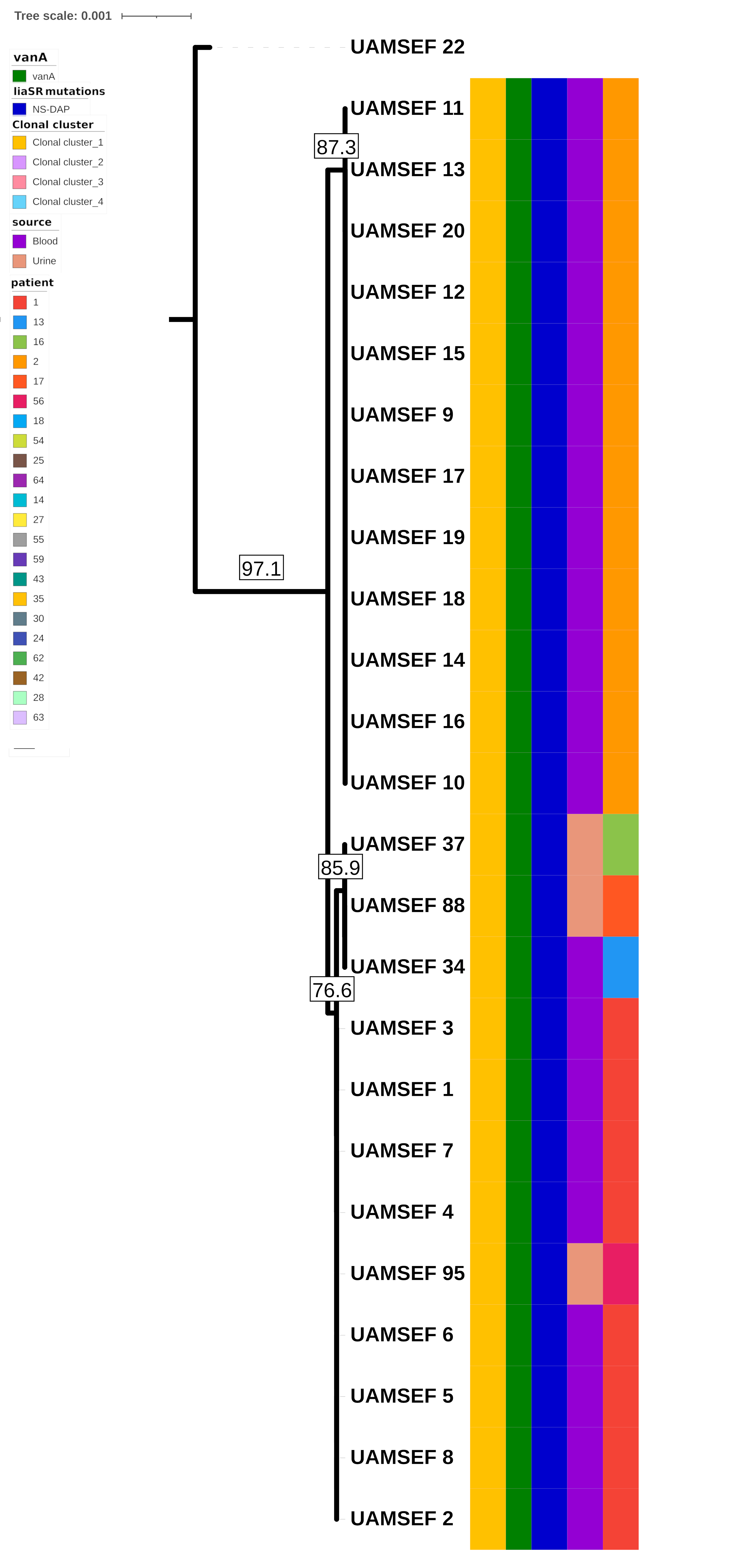

### Suppl. Fig4

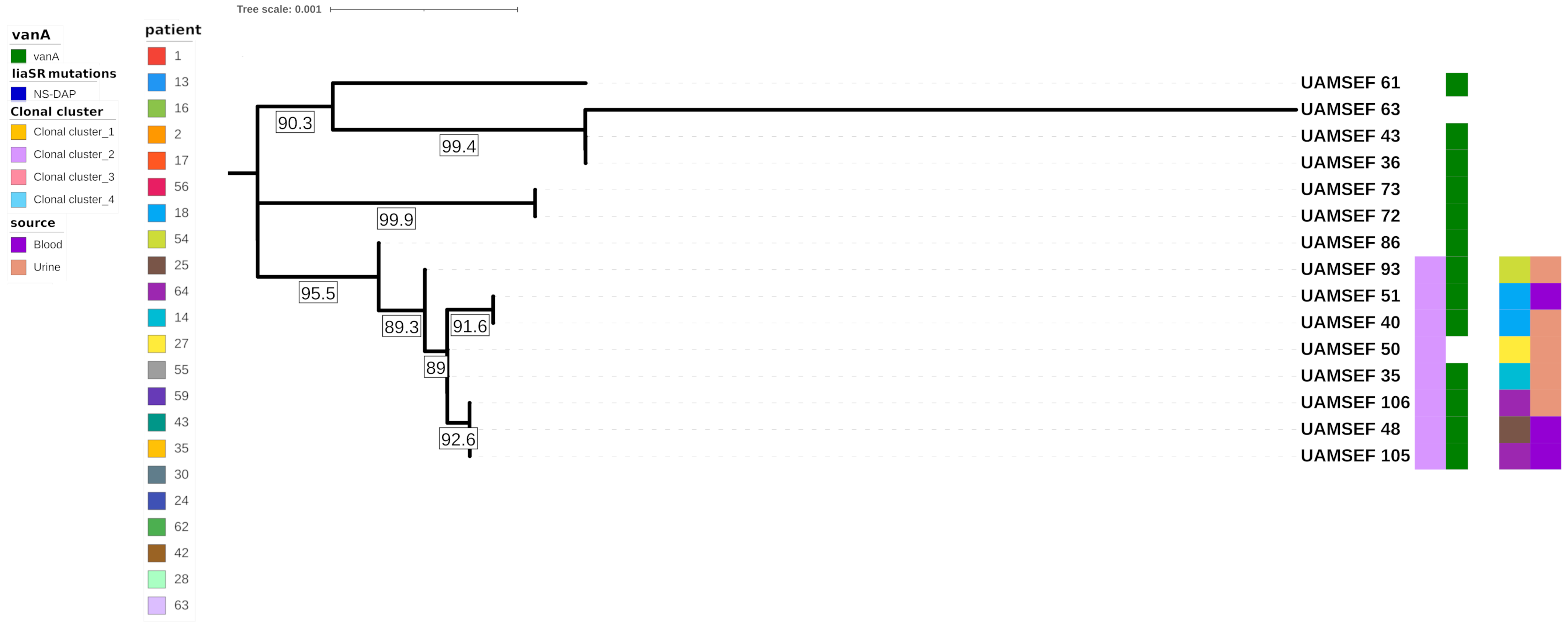

### Suppl. Fig5

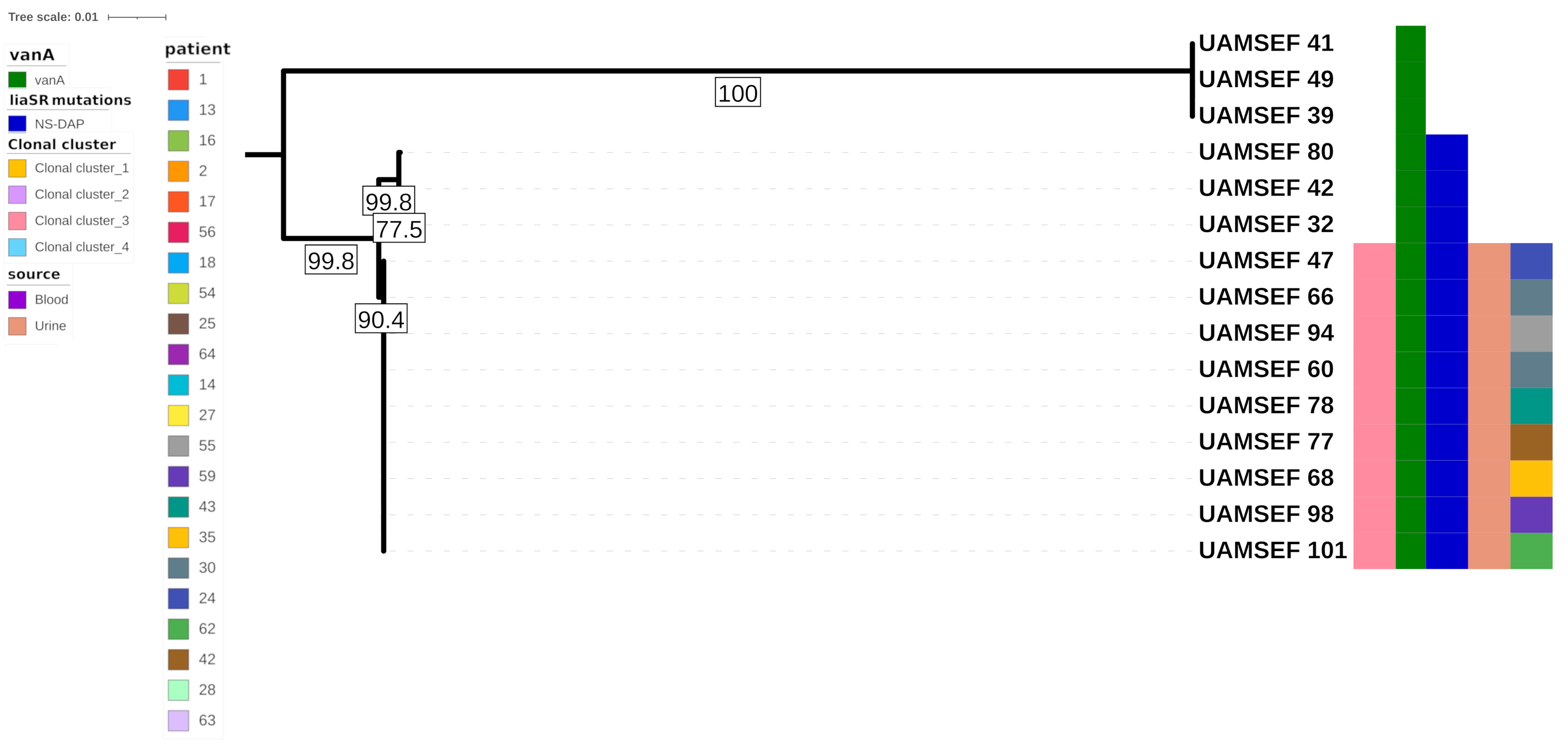

### Suppl. Fig6

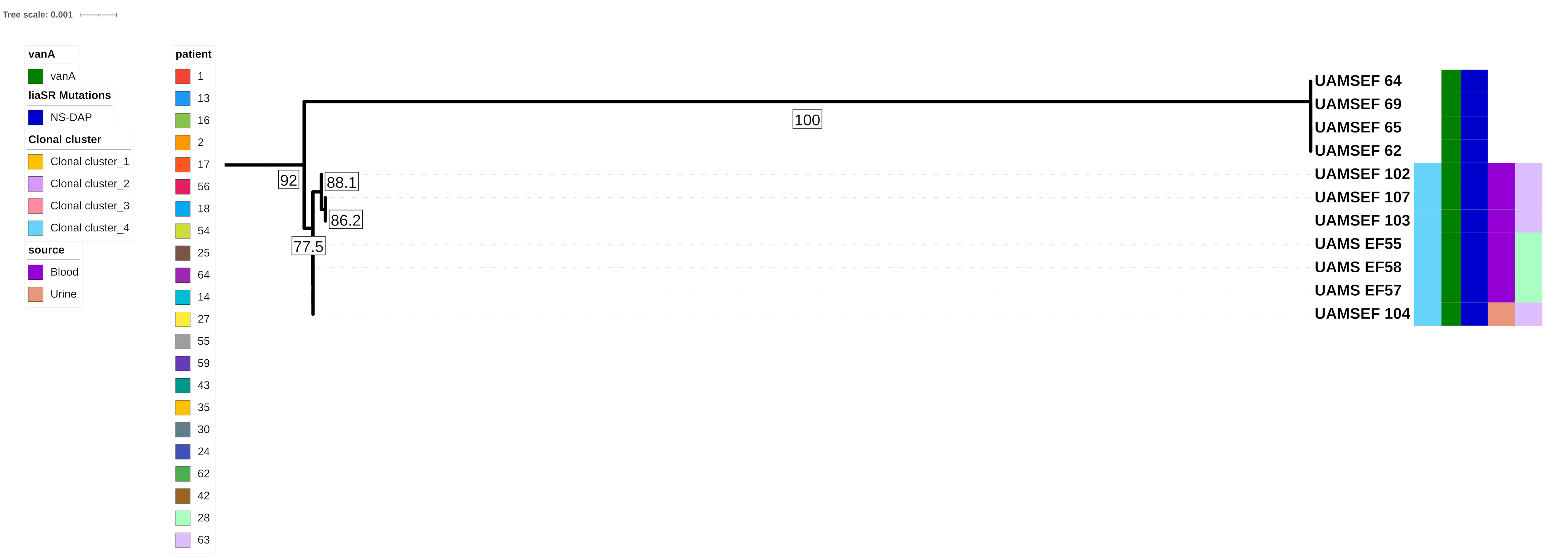

### Suppl. Figure 1

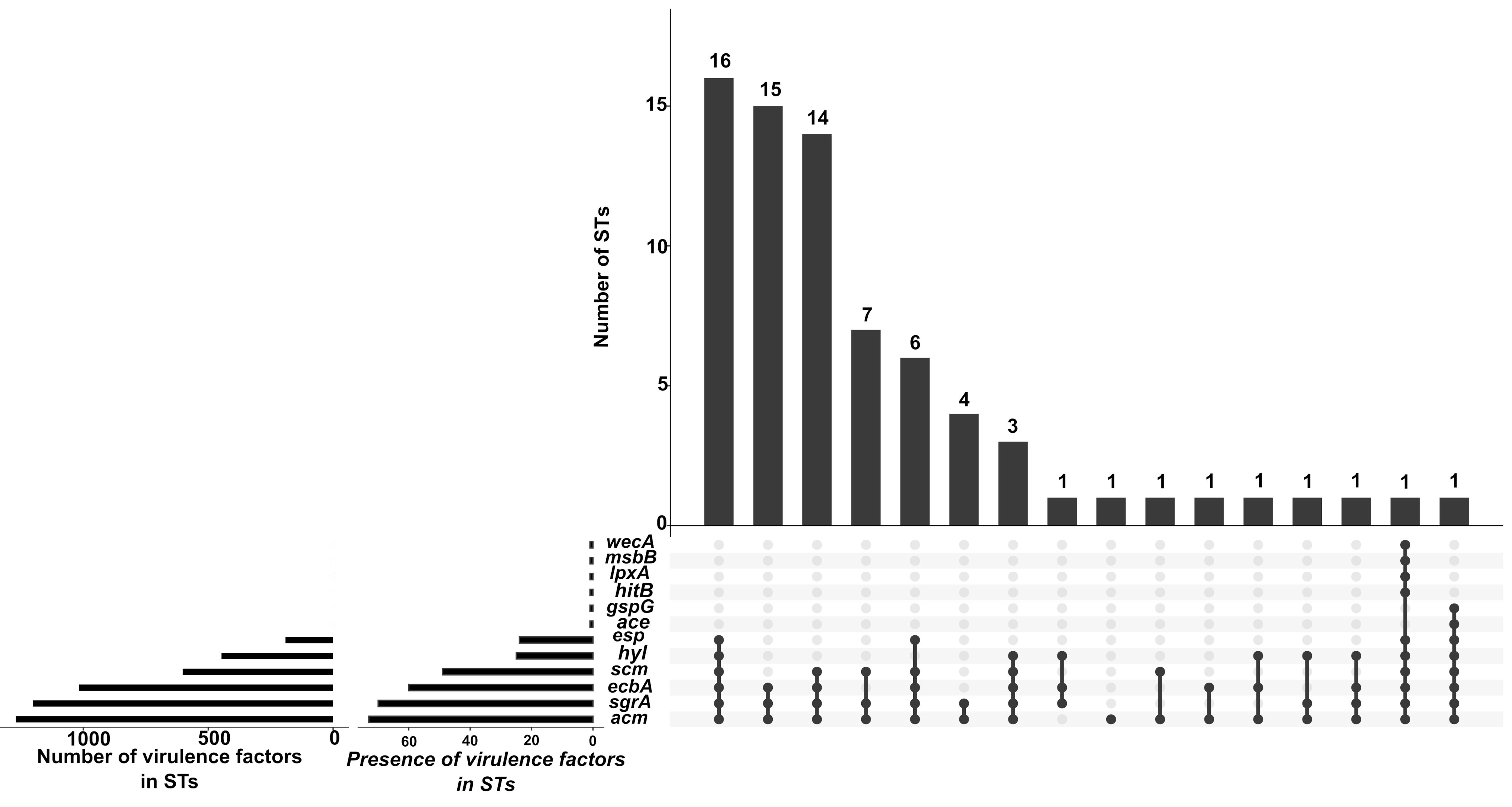
